# Descriptive Histomorphology of Palatine Tonsils Among Children Undergoing Tonsillectomy at a Zonal Referral Hospital in Dar es Salaam, Tanzania

**DOI:** 10.64898/2026.11.18.26368385

**Authors:** Johanes Joseph Kihanda, Noel Moses Nnyiti, Patrick Abrahams Karua, Denis A. Russa

**Affiliations:** Department of Anatomy, School of Medicine, Rabininsia University; Department of Otorhinolaryngology, School of Medicine, Rabininsia University; Otorhinolaryngology Unit, Department of Surgery, Rabininsia Memorial Hospital; Department of Anatomy, School of Biomedical Sciences, Muhimbili University of Health and Allied Sciences

## Abstract

**Background:** Palatine tonsils play a critical role in paediatric immune function, with their microarchitecture influencing susceptibility to disease. Despite the burden of tonsillar diseases in Tanzania, limited data exist on local histomorphological patterns that may underpin disease mechanisms. This study was conducted to evaluate the histomorphological characteristics of palatine tonsils in children undergoing tonsillectomy at a zonal referral hospital in Dar es Salaam.

**Materials and Methods:** This descriptive cross-sectional study analysed 86 tonsillectomy specimens collected from children aged ≤15 years between March and May 2025. Gross morphology (weight, surface, consistency) and histological features (epithelial thickness, crypt density, and high endothelial venule [HEV] density) were assessed using H&E staining and digital image analysis. Statistical analysis was performed using SPSS v20, with significance set at *p* < 0.05.

**Results:** Tonsillar hypertrophy was the most common surgical indication (54.7%). Heavier tonsils and smooth, soft surfaces were significantly associated with hypertrophy (*p* < 0.001), whereas recurrent tonsillitis correlated with irregular, firm tonsils. Surface epithelium was thicker than crypt epithelium (14 ± 3 vs. 12 ± 2 cell layers) and HEV density was higher in the surface epithelium (2.01 ± 0.63 vs. 1.55 ± 0.58/high power field). No statistically significant differences in epithelial thickness, crypt number, or HEV distribution were observed between clinical groups.

**Conclusion:** The study demonstrated conserved histological features across clinical groups with subtle tissue remodelling in disease states, suggesting that immunological and molecular responses may underlie the mechanisms of tonsillar disease in this population.

## INTRODUCTION

The palatine tonsils are paired lymphoid organs of Waldeyer’s ring that play a crucial role in mucosal immunity, particularly during early childhood ^1^. They are covered by stratified squamous epithelium which invaginates to form crypts, thereby increasing the surface area for antigen capture ^2^. Within the crypt epithelium, lymphoepithelial interactions facilitate direct contact between antigens and immune cells. The underlying lymphoid tissue is organized into follicles with germinal centres that support B-cell maturation, while the interfollicular areas harbour T-lymphocytes ^3^. High endothelial venules (HEVs) are prominent in these regions, serving as specialized entry sites for circulating lymphocytes, thus maintaining active immune surveillance^4^. Together, these structural features underline the palatine tonsils’ function as the first line of defence against inhaled and ingested pathogens ^5^.

Tonsillar diseases are a significant global health burden, with recurrent tonsillitis and hypertrophy being common indications for tonsillectomy ^6^. It is estimated that between 2% and 5% of outpatient visits worldwide are attributable to recurrent sore throat, and in children, the prevalence rises to 20–30% ^7^. In Tanzania, the considerable impact of tonsillar pathology on child health is reflected on the tonsillectomy rate. 56% of children attending otorhinolaryngology clinics undergo a tonsillectomy for recurrent tonsillitis or hypertrophy ^8^.

Research on tonsillar diseases in Tanzania has focused primarily on microbial colonization of tonsillar tissue, identifying bacterial pathogens associated with recurrent tonsillitis and their surface and core colonization patterns ^9,10^. Histomorphological aspects of the tonsils, which underlie their immune and pathological functions, remain poorly described in this context. Such knowledge is essential for understanding disease mechanisms, guiding clinical practice, and establishing local reference data. The present study was therefore conducted to describe the histomorphology of palatine tonsils among children undergoing tonsillectomy at a zonal referral hospital in Dar es Salaam, Tanzania. By providing baseline data on tonsillar structure in this population, the study aimed to contribute to improved understanding of tonsillar diseases and their clinical implications.

## MATERIALS AND METHODS

### Study Design, Setting and Duration

This was a descriptive cross-sectional study conducted at Rabininsia Memorial Hospital in Dar es Salaam between March 2025 and May 2025.

### Study Population

The study population included all children aged 15 years or less scheduled for tonsillectomy at Rabininsia Memorial Hospital for either recurrent tonsillitis or tonsillar hypertrophy. Informed consent was sought from the legal guardians of the children.

### Sampling Procedure

The participants were selected through convenience sampling method and enrolled until the sample size was achieved.

### Data Collection and Analysis

Collected specimens were examined macroscopically for surface texture and consistency before being fixed in 10% formalin. After fixation, they were transported to Muhimbili University of Health and Allied Sciences (MUHAS) histology laboratory. They were then weighed on an electronic scale. Parallel cross-sections were made and subsequently processed, embedded in paraffin and histological cross-sections of about 3 micrometres in thickness were made, stained with haematoxylin-eosin (HE) technique according to Bancroft for specimen evaluation. Three random fields of both surface and crypt epithelia were selected under a 40× objective lens, photographed and analysed using ImageJ software. Epithelial thickness was measured by counting cell layers, and HEVs were identified as vessels with at least one cuboidal endothelial cell. The crypt number was measured per mm^2^ using the objective lens magnification and ocular field number using the formula Field Diameter (mm) = (Ocular Field Number/Objective Lens Magnification), given the ocular field number of our microscope 20, and 10X objective lens, the diameter of our field of view was 2mm, giving a circular area of 3.14 mm^2^. Three random fields were examined, and mean crypt density was calculated.

### Statistics

Statistical analysis was performed using SPSS version 20 for Windows. Quantitative variables were expressed as mean ± standard deviation (SD) across surgical indication groups, and differences between groups were assessed using Student’s *t*-test. Qualitative variables were presented as frequencies and percentages, and associations were tested using the chi-square test. A *p*-value < 0.05 was considered statistically significant at the 95% confidence level.

### Ethical Considerations and Consent

Ethical approval for the study was obtained from the Institutional Review Board (IRB) at Muhimbili University of Health and Allied Sciences (MUHAS). Furthermore, permission to carry out the research was secured from the hospital authority of Rabininsia Memorial Hospital. The study complied with ethical principles in medical research, ensuring no harm to participants, promoting justice, and guaranteeing benefits without unnecessary risks. Guardians of the patients were given an informed consent form, which included the option to opt out of the study at any stage. Participant confidentiality was maintained rigorously throughout the research process.

## RESULTS

### Sociodemographic Characteristics

A total of 86 children scheduled for tonsillectomy were included in this study. The majority were male 55 (64%), while females accounted for 31 cases (36%). The predominant age group was 4– 6 years, representing more than half of the study population 46 (53.5%), whereas 20 children (23.3%) were younger than 4 years and another 20 (23.3%) were older than 6 years. Tonsillar hypertrophy was the most common indication for surgery, observed in 47 children (54.7%), while 39 children (45.3%) underwent tonsillectomy due to recurrent tonsillitis. (Table 1)

**Table 1:** Sociodemographic characteristics of patients, their surgical indication for tonsillectomy and gross morphological characteristics of their tonsils.

| **Variable** | **Frequency** | **Percentage (%)** |
|---|---|---|
| **Sex** |  |  |
| Male | 55 | 64 |
| Female | 31 | 36 |
| TOTAL | 86 | 100 |
| **Age (Years)** |  |  |
| 1-3 | 31 | 36 |
| 4-6 | 46 | 53.5 |
| Above 6 | 9 | 10.5 |
| TOTAL | 86 | 100 |
| **Indication for tonsillectomy** |  |  |
| Tonsillar Hypertrophy | 47 | 54.7 |
| Recurrent Tonsillitis | 39 | 45.3 |
| TOTAL | 86 | 100 |
| **Weight (Grams)** |  |  |
| I | 27 | 31.4 |
| II | 43 | 50 |
| III | 16 | 18.6 |
| TOTAL | 86 | 100 |
| **Mean ± SD** |  | **2.43 ± 0.09 grams** |
| **Surface** |  |  |
| Smooth | 47 | 54.7 |
| Irregular | 39 | 45.3 |
| TOTAL | 86 | 100 |
| **Consistency** |  |  |
| Soft | 41 | 47.7 |
| Firm | 45 | 52.3 |
| TOTAL | 86 | 100 |

### Gross Morphological Characteristics

Tonsil weight ranged from 0.75 grams to 5.44 grams with mean weight of 2.43 ± 0.09 grams. For descriptive purposes, the weights were categorized into categories; I (0.01-2 grams), II (2.01-3 grams) and III (Above 3 grams). The largest proportion of tonsils fell in category II, 43 (50%). On gross examination, the surface of the excised tonsils appeared either smooth or irregular. Out of the total 86 tonsils assessed, 47 (54.7%) had a smooth surface, while 39 (45.3%) exhibited an irregular surface. Assessment of tonsil consistency revealed that 41 (47.7%) of the tonsils were soft, whereas 45 (52.3%) were firm on palpation. (Table 1).

### Gross Morphological Characteristics according to Surgical Indication

Tonsil weight was significantly associated with indication for surgery (*p* < 0.001), with hypertrophy cases demonstrating heavier tonsils compared to recurrent tonsillitis. Additional gross features showed similar patterns: smooth surfaces were mostly seen in hypertrophy cases (78.7%), irregular surfaces most in recurrent tonsillitis (74.4%). Likewise, soft consistency was more frequent in hypertrophy (85.4%), whereas firm consistency was strongly associated with recurrent tonsillitis (73.3%). These associations were highly significant (*p* < 0.001). (Table 2).

**Table 2:** Gross morphology by surgical indication.

| **Parameter** |  | **Tonsillar Hypertrophy<br>N (%)** | **Recurrent Tonsillitis<br>N (%)** | **Total** | **Significance test** |
|---|---|---|---|---|---|
| Weight | I | 7 (25.9) | 20 (74.1) | 27 (100) | Chi-Square = 16.5419 P-value < 0.001 |
|  | II | 26 (60.5) | 17 (39.5) | 43 (100) |  |
|  | III | 14 (87.5) | 2 (12.5) | 16 (100) |  |
|  | Total | 47 (54.7) | 39 (45.3) | 86 (100) |  |
| Surface | Smooth | 37 (78.7) | 10 (21.3) | 47 (100) | Chi-Square = 24.833 P-value < 0.001 |
|  | Irregular | 10 (25.6) | 29 (74.4) | 39 (100) |  |
|  | Total | 47 (54.7) | 39 (45.3) | 86 (100) |  |
| Consistency | Soft | 35 (85.4) | 6 (14.6) | 41 (100) | Chi-Square = 29.826 P-value < 0.001 |
|  | Firm | 12 (26.7) | 33 (73.3) | 45 (100) |  |
|  | Total | 47 (54.7) | 39 (45.3) | 86 (100) |  |

### Histological Characteristics

The surface epithelium was found to be slightly thicker than the crypt epithelium (14 ± 3 cells vs. 12 ± 2 cells). Additionally, the density of high endothelial venules (HEVs) was higher in the surface epithelium (2.01 ± 0.63) per high power field compared to the crypt epithelium (1.55 ± 0.58) per high power field. There were no statistically significant differences in epithelial thickness or HEV distribution between patients with tonsillar hypertrophy and those with recurrent tonsillitis. The surface epithelium thickness was similar in both groups (14 ± 2 cells vs. 14 ± 3 cells, p = 0.862), as was crypt epithelium thickness (12 ± 2 cells vs. 11 ± 2 cells, p = 0.603). Likewise, HEV distribution in both the surface and crypt epithelia showed no significant variation between groups, with values of 2.01 ± 0.69 per high power field vs. 1.96 ± 0.95 per high power field (p = 0.471) and 1.53 ± 0.56 per high power field vs. 1.57 ± 0.61 per high power field (p = 0.703), respectively. The number of tonsillar crypts per mm^2^ appeared nearly the same in both groups studied. (Table 3).

**Figure 1:**
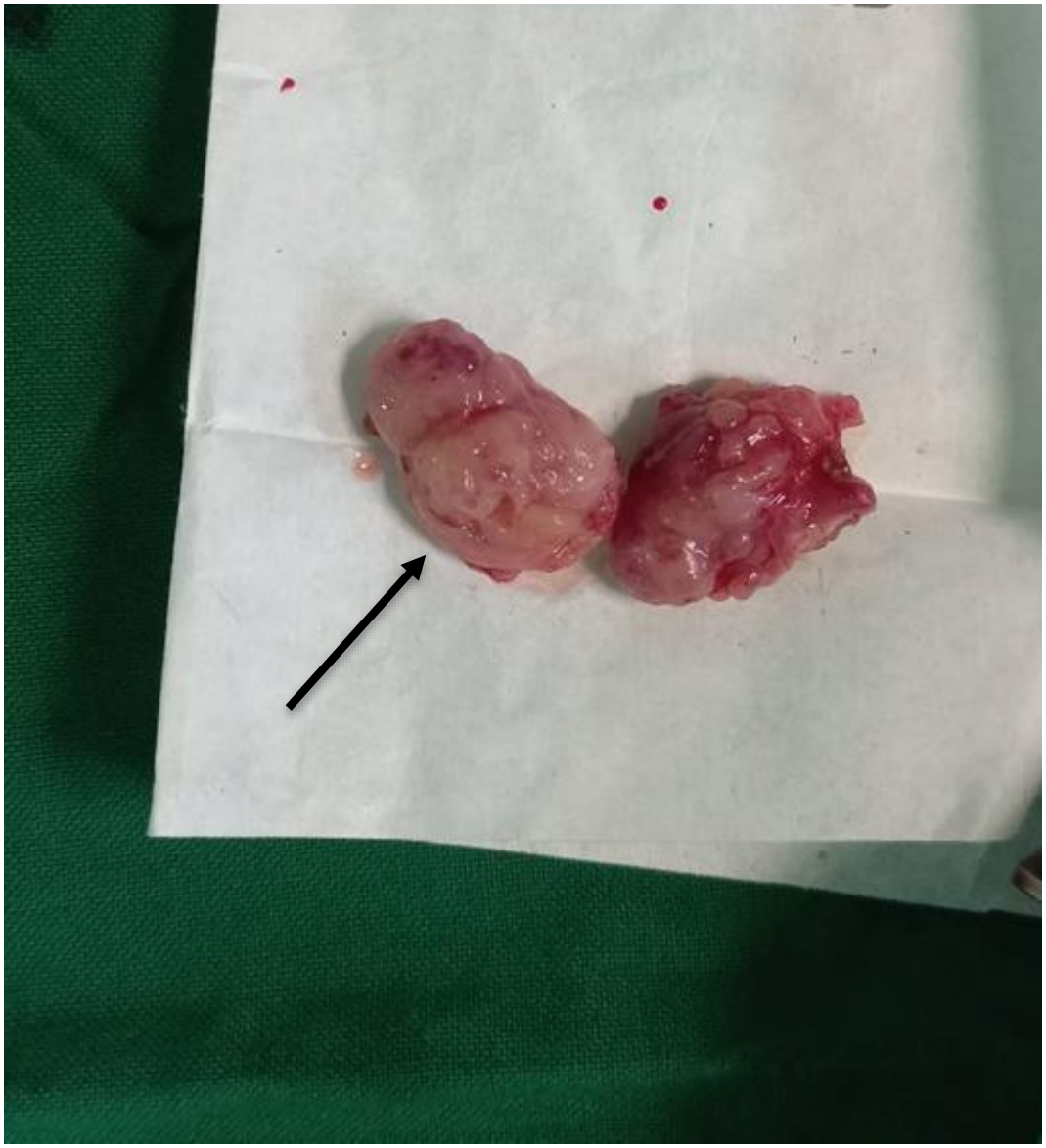
Palatine tonsils removed due to recurrent tonsillitis. Black arrow; The irregular texture of the oropharyngeal surface can be seen

**Figure 2:**
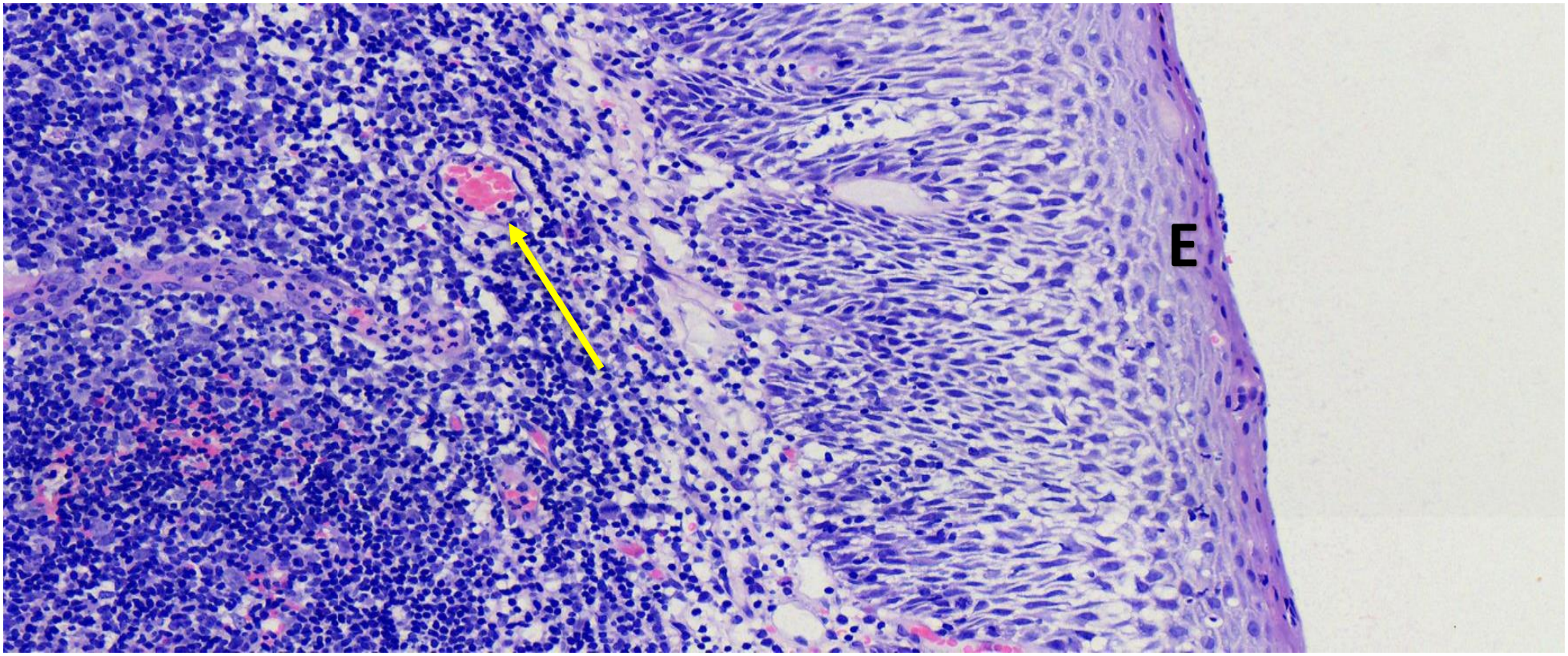
Histological section of palatine tonsil stained with Haematoxylin and Eosin (H&E) under 200X magnification. E = Surface Epithelium, Yellow arrow: High Endothelial Venule

**Figure 3:**
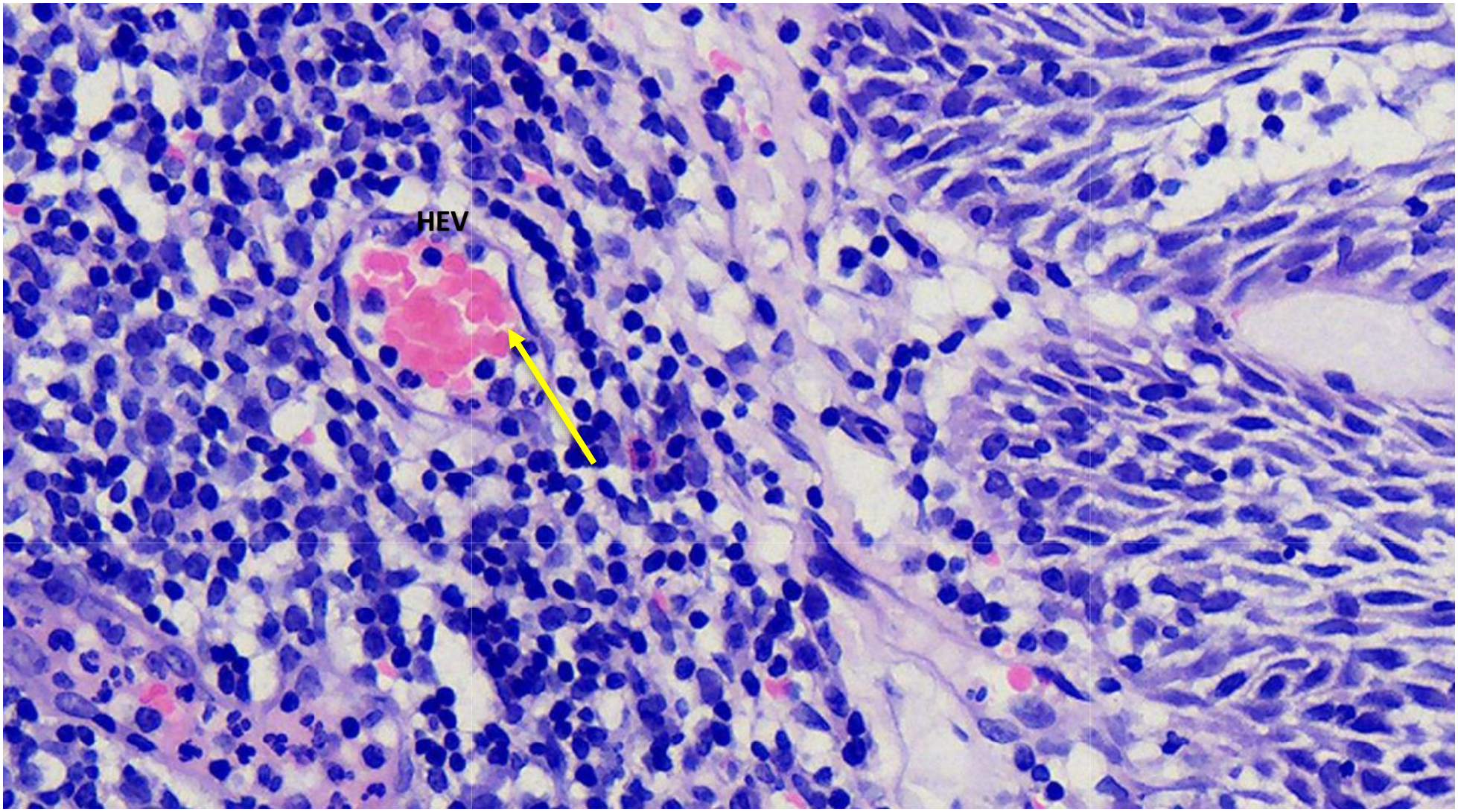
Histological section of palatine tonsil stained with Haematoxylin and Eosin (H&E) under 400X magnification to show the high endothelial venule. HEV = High Endothelial Venule, Yellow arrow: Cuboidal endothelial cell of HEV

**Figure 4:**
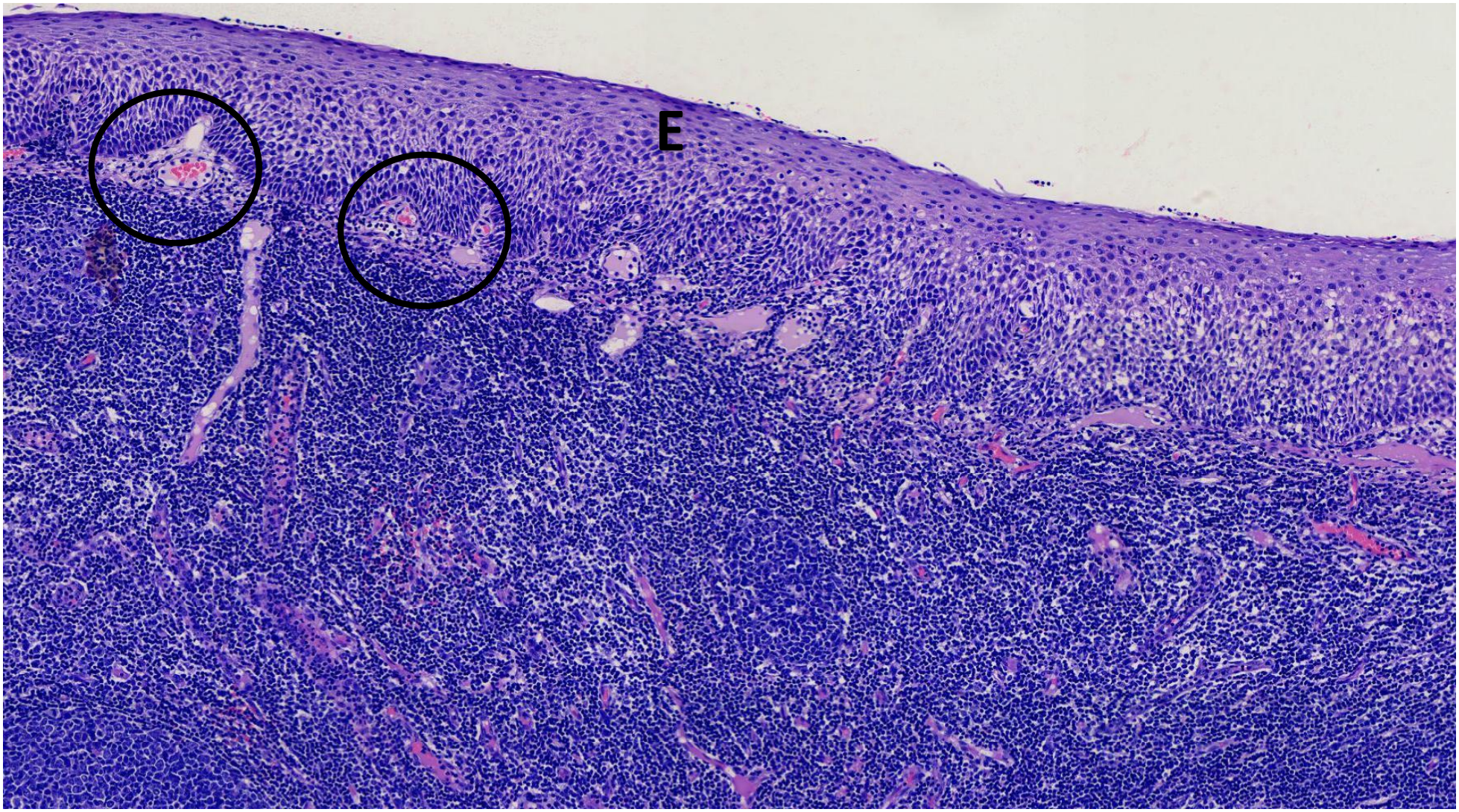
Histological section of palatine tonsil stained with Haematoxylin and Eosin (H&E) under 100X magnification. E = Stratified squamous epithelium, Black circles: Invagination of the epithelium

**Table 3:**
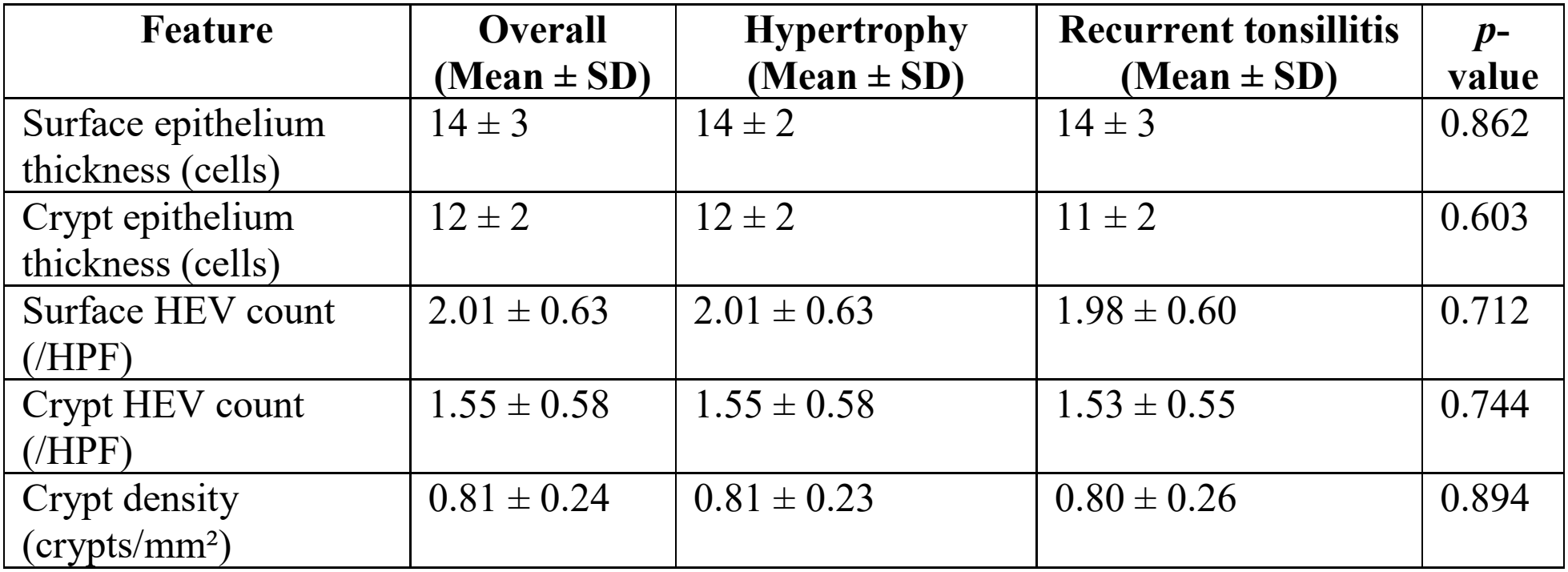
Histological features.

| **Feature** | **Overall (Mean ± SD)** | **Hypertrophy (Mean ± SD)** | **Recurrent tonsillitis (Mean ± SD)** | ***p*-value** |
|---|---|---|---|---|
| Surface epithelium thickness (cells) | 14 ± 3 | 14 ± 2 | 14 ± 3 | 0.862 |
| Crypt epithelium thickness (cells) | 12 ± 2 | 12 ± 2 | 11 ± 2 | 0.603 |
| Surface HEV count (/HPF) | 2.01 ± 0.63 | 2.01 ± 0.63 | 1.98 ± 0.60 | 0.712 |
| Crypt HEV count (/HPF) | 1.55 ± 0.58 | 1.55 ± 0.58 | 1.53 ± 0.55 | 0.744 |
| Crypt density (crypts/mm<sup>2</sup>) | 0.81 ± 0.24 | 0.81 ± 0.23 | 0.80 ± 0.26 | 0.894 |

## DISCUSSION

This study aimed to characterize the gross and histomorphological features of palatine tonsils among children undergoing tonsillectomy at a zonal referral hospital in Dar es Salaam, Tanzania. By examining gross and histological features such as tonsil weight, surface texture, consistency, epithelial thickness, crypt density, and vascular distribution, the study aimed to provide insight into the structural attributes underlying tonsillar disease and establish baseline anatomical and histological profiles within the local paediatric population. The demographic profile of the study population mirrors trends observed in similar paediatric studies. Males comprised 64% of the participants, and the age group four to six years represented the largest proportion (53.5%). These findings align with global patterns indicating increased lymphoid tissue activity in early childhood, especially between ages 3 and 7, often resulting in tonsillar pathology ^6^. Upper airway obstruction (UAO), primarily related to tonsillar hypertrophy, was the leading indication for surgery (54.7%), corroborating the global and local shift from infectious indications toward obstructive symptoms as primary drivers for tonsillectomy ^8,11,12^.

The mean tonsil weight was 2.43 ± 0.09g, with half of the specimens (50%) falling within the 2.01– 3g range. Males and children older than 6 years exhibited a higher frequency of heavier tonsils (category III), though this association was not statistically significant. However, there was a highly significant relationship between tonsil weight and surgical indication (p < 0.001), with heavier tonsils associated with hypertrophy and lighter tonsils linked to recurrent tonsillitis. This finding substantiates clinical expectations: hypertrophic tonsils are more likely to undergo hyperplasia, resulting in increased mass, whereas recurrently infected tonsils may be chronically inflamed and fibrotic but not necessarily enlarged ^13^. Importantly, surface texture and consistency demonstrated significant diagnostic potential. Tonsils with a smooth surface and soft consistency were predominantly seen in cases of tonsillar hypertrophy (78.7% and 85.4% respectively). Conversely, irregular surfaces and firm consistency were strongly associated with recurrent tonsillitis (74.4% and 73.3%, respectively), with both relationships reaching high statistical significance (p < 0.001). These distinctions may reflect underlying histological remodelling: chronic infection can lead to fibrosis, crypt scarring, and surface irregularities, while hypertrophy may be more associated with lymphoid hyperplasia and minimal disruption of surface architecture ^14^. These gross features may offer surgeons additional intraoperative clues to the underlying pathology and could support more informed clinical decision-making.

Microscopically, the study focused on three key features: epithelial thickness, HEV (high endothelial venule) density, and crypt density. Across the entire sample, the surface epithelium was thicker than the crypt epithelium (14 ± 3 vs. 12 ± 2 cells), consistent with previous literature suggesting the surface is more robust to withstand mechanical and antigenic exposure ^15^. HEV density was also higher in the surface epithelium (2.01 ± 0.63/HPF) than in crypt regions (1.55 ± 0.58/HPF), possibly indicating more active lymphocyte trafficking at the surface level in this cohort ^16^. Interestingly, while gross features differed markedly between clinical groups, histological parameters did not vary significantly between tonsillar hypertrophy and recurrent tonsillitis. For example, surface epithelial thickness was identical (14 ± 2 vs. 14 ± 3 cells; p = 0.862), and crypt density showed negligible variation (0.81 vs. 0.80 crypts/mm^2^; p = 0.894). These results align with studies suggesting that while chronic inflammation may lead to subtle microarchitectural remodelling, the foundational structure of the tonsil, remains largely preserved across different clinical conditions ^15,17^. The lack of significant difference in HEV distribution between the two groups (p-values 0.471 and 0.703 for surface and crypt regions respectively) diverges from some international findings. Prior research, such as that by Tabou et al. and Jović et al., reported increased HEV density in the crypts of tonsils affected by recurrent tonsillitis, reflecting enhanced lymphocyte trafficking ^18,19^. The discrepancy may be attributed to methodological differences: whereas those studies employed immunohistochemical staining (e.g., CD34 or CD146) for precise HEV identification, the current study relied on H&E staining, which may under-represent subtle vascular changes. Another potential explanation lies in the local population’s unique immunological profile or environmental exposures, which may influence how tonsillar tissue responds to chronic stimuli ^20,21^. The similar histological architecture across groups may also support a model where functional or molecular responses rather than structural ones drive the clinical manifestation of disease.

### Study Limitations

This study was conducted at a single zonal referral hospital which may limit the generalizability of findings to broader paediatric populations across Tanzania. The study also relied solely on H&E staining for identifying histological features such as high endothelial venules which is potentially less sensitive than immunohistochemical techniques which would have provided more specific and sensitive identification of these features.

## CONCLUSION

This study reveals that while histological architecture of the palatine tonsils is largely conserved between hypertrophy and recurrent tonsillitis, gross morphological characteristics demonstrated strong associations with surgical indication.

## RECOMMENDATIONS

These findings highlight the diagnostic value of meticulous gross examination and the conserved histological features offer limited discriminatory power unless supplemented by advanced staining or molecular tools. The results support the pursuit of refined functional and molecular studies to better understand the underlying mechanisms of tonsillar disease in paediatric populations and inform future clinical and research priorities in resource limited settings.

## Data Availability

All data produced in the present study are available upon reasonable request to the authors

## Acknowledgements

I would like to express my heartfelt appreciation to my supervisors for their consistent guidance, support and constructive feedback which were instrumental in shaping the direction of this research. I am also grateful to the laboratory technicians at MUHAS for their technical assistance and dedication during the laboratory procedures. Sincere thanks are extended to Rabininsia Memorial Hospital for their cooperation during specimen collection, as well as to the hospital staff whose support was vital to the study’s progress.

